# Perception and awareness of COVID-19 among health science students and staff of Kuwait University: An online cross-sectional study

**DOI:** 10.1101/2020.12.26.20248891

**Authors:** Walid Q. Alali, Wadha AlFouzan, Dhuha Aljimi, Haya Al-Tawalah, Khalid A. Kheirallah, Getnet Yimer

## Abstract

**Background:** Coronavirus disease 2019 (COVID-19) pandemic is unprecedented. Health science students are the future frontliners to fight pandemics. Awareness and perception toward COVID-19 among health science students and staff at Kuwait University was assessed.

**Methods:** Between June and July 2020, an online questionnaire was distributed to all students and staff at HCS. The questionnaire was divided into six sections: socio-demography, risk and awareness, preparedness and prevention, source of information, policies, and social stigma.

**Results:** A total of 592 students and 162 staff completed the questionnaire. The prevalence of self-reported chronic condition among students and staff was 14.0% and 19.1%, respectively. Moreover, self-reported COVID-19 prevalence among students and staff was 2.7% and 1.2%, respectively. Interestingly, 54% of students and 38.3% of staff reported that they knew someone within their immediate social environment who have been/are infected with SARS-CoV-2. Among students, 92.4% wore face mask in indoor places (outside of their home) often/all the time compared to wearing it outdoors (69.3%); whereas, for staff, it was more common to wear it outdoor than in indoor places (75.9% vs. 81.5%). Both students and staff showed greatest deal of trust was in official government press release and consultation with healthcare workers.

Willingness to take COVID-19 vaccine was indicated by 50% of students strongly agreed and an additional 25.8% agreed to taking it. Interest vaccine uptake was lower among staff (28.4% and 34.6% strongly agreed or agreed, respectively). Participants strongly agreed or agreed (72.5% and 19.6% of students as well as 68.5% and 22.2% of staff) that wearing face mask in public should be obligatory. More than 18% of students and staff indicated that they would avoid contact with COVID-19 infected people.

**Conclusions:** Responses of students and staff were mostly similar and showed that they follow precautionary measures to control spread of COVID-19, understand the viral transmission risk, and willing to raise awareness to reduce social stigma.

## Introduction

The coronavirus disease 2019 (COVID-19) caused by the severe acute respiratory syndrome coronavirus 2 (SARS-CoV-2) continues to be a global public health threat [1]. Without an effective treatment or vaccine, the virus is expected to stay for long period and infect more people.

The global health crisis of COVID-19 has grabbed the attention of the mass media around the world [2]. Most of the public health messages have been focused on the control of transmission risk. Many of these messages are related to social distancing, frequent and proper hand washing, stay at home, and avoid crowded places. However, the pandemic of information has caused a great deal of anxiety, fear, excessive worry, and confusion among people worldwide. Additionally, the rapidly evolving and changing knowledge about the disease have created a cloud of misinformation and miscommunication among people in different communities [3-5].

Kuwait went through a period of partial and full lockdowns between May and August 2020 following the large increase in cases and deaths in March and April 2020 [6]. School closure (including universities) was one of the first restrictions applied at the beginning of the epidemic (March 2020) in Kuwait and continued during the study period. Many countries worldwide have gone through similar school closure and continue to have it in effect [7].

The COVID-19 pandemic is unprecedented, and no other disease has dominated the human population for the past 100 years. Human behavior is partially driven by the knowledge and perception gained through exposure to information from various sources such as formal education, media, family, friends and colleagues [8]. In Kuwait, people receive information about COVID-19 from different sources including the Ministry of Health and official state TV, World health organization, expert and non-expert opinions on social media, as well as friends and colleagues. Therefore, this study was focused on assessing perception and awareness on matters related to transmission, control and prevention of COVID-19; sources of information; and social stigma. We believe the study is important for two main reasons: 1) health education and raising awareness about the disease among university students and staff as well as the larger community, and 2) findings from this survey can aid the policy makers in their decision making to control the epidemic in Kuwait and apply social policies in relation to this population.

There are number of published studies that assessed the epidemiology, pathogenesis, virology, and treatment of COVID-19. However, there are limited studies assessing the COVID-19 perception, attitude, and awareness in the academic community [9]. The academic community is an important segment of the society and represent students and staff in higher education institutions. Students in the health science fields (e.g., medicine, dentistry, pharmacy and public health) will be the next generation of health leaders; hence, assessing their perception in relation to the current pandemic is expected to yield useful outcomes.

Kuwait University is the state national university with a student population of about 35,000 students. The Health Science Center (HSC) of Kuwait University is composed of five faculties including Medicine, Dentistry, Pharmacy, Public Health, and Allied Health. They represent a more uniformed population compared to the rest of the university’s population. Furthermore, many of the HSC staff and students have been involved with COVID-19 related activities including volunteering in health facilities, raising awareness about the disease and pursue research opportunities. The goal of this study was to describe the awareness and perception on matters related to control and prevention of COVID-19, source of information and social stigma among students and staff of HSC at Kuwait University.

## Methods

### Study population

A cross-sectional survey study was conducted between June and July 2020 on students and staff at HCS of Kuwait University. Our target populations were: all undergraduate and postgraduate students enrolled in the five faculties (Medicine, Dentistry, Pharmacy, Public Health and Allied Health) of HSC and all the staff (faculty members, teaching assistants, and supporting staff) employed by HSC. At the time of the study, there was approximately 2,000 undergraduate students and about 110 graduate students (Master and PhD programs). In addition, there was approximately 450 staff employed by HSC at the time of the study.

### Data collection tool

Data collection occurred via online questionnaire distributed to students and staff. During the study period, the university was closed, and no classes were held on campus nor through distance learning.

The questionnaire was developed based on validated survey tools provided by World Health Organization [10] and that used by Khasawneh et al. [11]. Nonetheless, the questionnaire was adapted to our target population. Non-relevant or redundant questions were excluded from both survey tools and those that met the objectives of this study were included. Some of the questions taken from the previous survey tools were modified such as changes in the number of multiple-choice answers per question as well as rewording the questions to improve clarity.

The questionnaire was divided into six sections: socio-demography, risk and awareness, preparedness and prevention, source of information, policies, and social stigma. Socio-demography questions included questions about gender, academic year, faculty, and presence of chronic illness. Additional questions for staff included job classification and age. Risk and awareness section included questions about history of COVID-19 infection, risk of infection in the future and infection severity, awareness of consequences of infection as well as awareness of availability of treatment or vaccine for COVID-19. Preparedness and prevention section included questions about probability of avoiding infection with SARS-CoV-2, prevention measures taken to avoid infection measures as frequency based on the participants response to one of the following options: most or times, rarely or sometimes, and never. Additional question on participants’ opinion on attending classes or working on-campus vs. online learning methods. The source of information section had questions about main sources of acquiring information. These included Television news stations, family and friends, healthcare workers, online search, social media, official government press releases, and celebrities. Policies section included questions about COVID-19 vaccine timeline and willingness to uptake the vaccine and treatment.

Furthermore, opinion of participants regarding the following: mandatory quarantine of COVID-19 contacts, wearing face mask in public places, legal punishment of who is COVID-19 infected and knowingly socialize with others and those spread share/spread falsified information about COVID-19. Social stigma section included questions about: participants’ reaction to knowing someone COVID-19 positive, blaming the person of who got the infection, what information the participants would like to know about those infected, community reaction to those infected (such as being socially rejected, denied medical care, job loss), and ways to reduce social stigma.

Two questionnaires were developed; one administered to students and another to the staff. Both questionnaires were very similar with only differences in the first section (i.e., socio-demography). The questionnaires were provided in English since the students and staff are familiar with this language.

The questionnaires were distributed and administered to the participants online via Google Forms (www.google.com/forms). The questionnaires were sent via an email listserv to all students and staff within HSC. Additionally, student representatives at each faculty distributed the questionnaire to class groups via WhatsApp. Each of the six sections was composed of several questions (*supplementary file A*). The questionnaires were validated and pilot tested on 10 students and 10 staff prior to administration to all participants. Multiple reminders were sent to all participants to increase response rate.

### Statistical analysis

Data were exported from Google Forms to an Excel spreadsheet for data management. The analyses were conducted in STATA statistical software ver. 15.1 (College Station, Texas). A descriptive analysis using cross tabulation of frequency and percentage of variables was conducted. Students were categorized into two groups: younger students (freshmen and junior student [years 1-3]) and older students (senior [years 4-7] and graduate students)) for the purpose of the univariate (chi-square) analysis in STATA. Alpha of 0.05 was used as the significance level.

## Results

### Demographics of participants

The demographics of students (n=592) and staff (n=162) are shown in Table 1. Most the participants were females (for both students and staff). Female students account for 75% to 90% of the student population depending on the academic year and the faculty. Participants were proportionally distributed to the population of their faculties with the largest faculties being Medicine and Allied Health. Only Medicine and Dentistry faculties have a seven-year academic program; whereas Pharmacy is 5-year program and both Allied Health and Public Health are 4-year programs. While the undergraduate students’ age ranged between 19 and 26 years; graduate students’ age was between 25 and 45 years. About 50% of the staff participants were faculty members. Most of the staff were older than 30 years with 24% of them over the age of 50 years.

**Table 1:** Sociodemographic of study participants.

| HSC Students (n=592) |  |  | HSC Staff (n=162) |  |  |
| --- | --- | --- | --- | --- | --- |
| Characteristic |  | No. (%) | Characteristic |  | No. (%) |
| Gender |  |  | Gender |  |  |
|  | Female | 545 (92.1) |  | Female | 124 (76.5) |
|  | Male | 47 (7.9) |  | Male | 38 (23.5) |
| Faculty |  |  | Faculty |  |  |
|  | Faculty of Allied Health | 135 (22.8) |  | Faculty of Allied Health | 24 (14.8) |
|  | Faculty of Dentistry | 57 (9.6) |  | Faculty of Dentistry | 15 (9.3) |
|  | Faculty of Medicine | 223 (37.7) |  | Faculty of Medicine | 80 (49.4) |
|  | Faculty of Pharmacy | 131 (22.1) |  | Faculty of Pharmacy | 21 (12.9) |
|  | Faculty of Public Health | 46 (7.8) |  | Faculty of Public Health | 22 (13.6) |
| Academic Year |  |  | Job Classification |  |  |
|  | First Year | 108 (18.2) |  | Faculty Member | 80 (49.4) |
|  | Second Year | 70 (11.8) |  | Teaching Assistant | 56 (34.6) |
|  | Third Year | 90 (15.2) |  | Supporting non-academic staff | 26 (16.1) |
|  | Fourth Year | 119 (20.1) | Age |  |  |
|  | Fifth Year | 68 (11.5) |  | 20 - 30 | 22 (13.6) |
|  | Sixth Year | 67 (11.3) |  | 31 - 40 | 60 (37.0) |
|  | Seventh Year | 46 (7.8) |  | 41 - 50 | 41 (25.3) |
|  | Graduate (Any Year) | 24 (4.1) |  | >50 | 39 (24.1) |

On chronic condition reporting, the prevalence of self-reported having any chronic condition among students and staff was 14.0% and 19.1%, respectively. The highest chronic condition among students was asthma (42.2%) and among the staff was hypertension (36.6%).

### Risk and awareness

The participants answered 10 questions related to SARS-CoV-2 risk of infection, risk of transmission and their information awareness about the virus. Based on participants’ self-reported SARS-CoV-2 infection (based on PCR confirmation test), the prevalence among students and staff was 2.7% and 1.2%, respectively. However, 85.0% and 86.4% of students and staff, respectively, reported that they have not been infected with the virus. Interestingly, 54.0% of students and 38.3% of the staff reported that they knew someone in their immediate social environment, such as relatives or friends, who have been/are infected with SARS-CoV-2.

When we asked the participants about their probability of getting infected with the virus in the near future, 3.9% of students responded as it was extremely unlikely; 23.3% indicated it is unlikely, but 5.5% responded it was likely. As for the staff, 5.6%, 21%, and 20.4% considered the probability to be extremely unlikely, unlikely, and likely, respectively. There was no significant (P = 0.374) relationship between the infection probability among staff participants and their age groups. Many of the students expressed that if they get infected with the virus, the disease will ‘not be severe’ (27.7%) or ‘slightly severe’, (24.6%). Similarly, many of the staff (20.4% and 21.6%) thought that if they get infected with the virus, the disease will ‘not be severe’ or slightly severe’, respectively.

The participant’s responses to questions regarding awareness related to proportion of COVID-19 patients that are likely to be admitted to hospital; if admitted to the hospital are likely to be admitted to the ICU; and if admitted to ICU will eventually die is shown in Figure 1. Most of the participants either strongly disagreed or disagreed with the following statements: 1) majority of COVID-19 patients need hospitalization; 2) those hospitalized require ICU admission; and 3) those admitted to ICU will eventually die (Figure 1). Older students were significantly (P <0.001) more likely to strongly disagree or disagree than younger students with the first two statements. No significant difference (P = 0.576) was detected between both student groups to the third statement. The faculty members were significantly more likely (P <0.001) to strongly disagree or disagree compared to teaching assistant and non-academic staff. No significant difference (P = 0.583) detected between staff job groups by the response to those admitted to ICU will eventually die.

**Figure 1.**
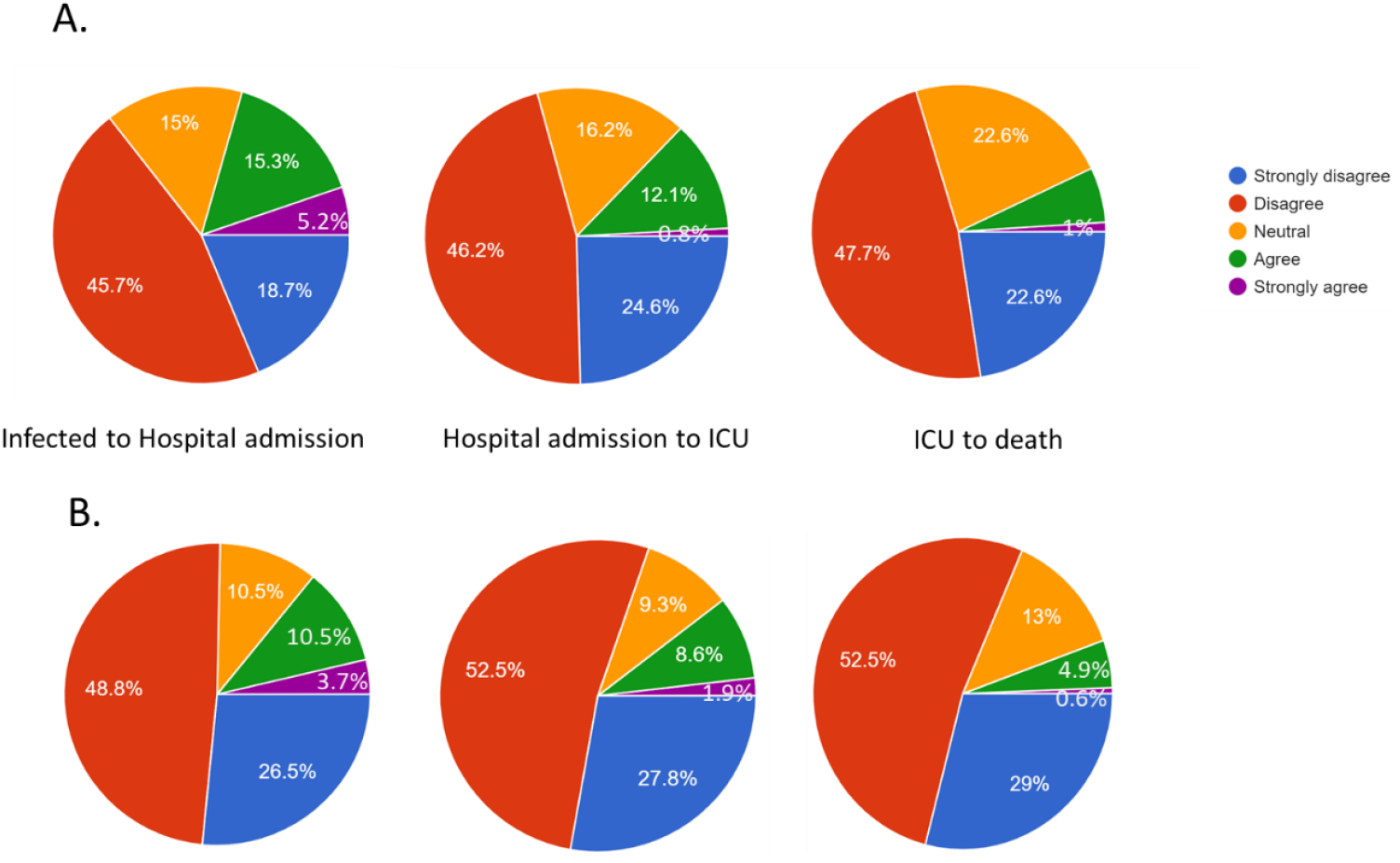
Participants awareness of consequences of COVID-19 patients* * Infected to hospital admission is based on the question: Do you agree or disagree that majority of SARS-CoV-2 infected people need admission to a hospital; Hospital to admission to ICU is based on the question: Do you agree or disagree that majority of SARS-CoV-2 patients who are admitted to the hospital require receiving care in the intensive care unit (ICU); and ICU to death is based on the question: Do you agree or disagree that majority of SARS-CoV-2 patients who are admitted to the ICU will eventually die. A: students and B: staff.

We asked the participants about their reaction to suspicion of being infected, nearly 52% of the students and 50% of staff indicated that they would immediately get tested for the virus and follow government isolation instructions (if tested positive); whereas, 28.2% and 36.4% of students and staff, respectively, would stay at home and taking precautionary measures so they do not spread the virus to others, and 19.6% and 13% of students and staff, respectively, would wait for few days until symptoms worsen before they get tested and/or seek medical care. On the topic of treatment or vaccine for COVID-19, most of the participants (80.6% of students and 85.8% of staff) indicated that, at the time of this study, there is no treatment or vaccine.

On the question of self-rate perception on how to prevent SARS-CoV-2 spread, most students rated themselves as having either very good knowledge (30.4%) or good knowledge (45.7%) with significantly (P <0.001) higher proportion among older students than younger students. On the same question, 43.8% and 37.7% of staff rated themselves as having very good knowledge and good knowledge, respectively, with significantly (P = 0.02) higher proportion among faculty members compared to other groups. In term of misconceptions, students either strongly disagreed (19.4%) or disagreed (21.8%) that SARS-CoV-2 was made in a laboratory then accidentally released. Similarly, 14.2% and 17.3% of staff either strongly disagrees or disagreed, respectively. Moreover, while most of students (30.2% strongly disagreed or 40.6% disagreed) that hot summer slowdown/stop the transmission of the virus, significantly (P <0.001) fewer staff strongly disagreed and disagreed (14.2% and 17.3%, respectively) with that statement.

### Precautionary measures to prevent infection with SARS-CoV-2

The participants answered 15 questions about precautionary measures for SARS-CoV-2 infection (Table 2). Among students, 92.4% wore face mask in indoor places (outside of their home) ‘often/all the time’ compared to wearing it outdoors (69.3%); whereas, for staff, it was more common to wear it outdoor than in indoor places (75.9% vs. 81.5%). Regular hand washing was more common than using disinfectants among both participant’s groups (Table 2). Avoiding crowded places, avoiding shaking hands, avoiding kissing, staying home as much as possible and following social distancing guidance was practiced ‘often/all the time’ by more than 80% of the participants. There was no significant difference (P>0.05) between staff than students with regard to these percentages. Nearly 50% of students did often clean/disinfect their phone, avoided eating outside of home, got sufficient sleep, and persuade people around them to follow precautionary guidance. This percentage was between 60-70% among staff (Table 2). Additionally, we asked the participants a question on how difficult/easy it is to avoid infection with the virus, 27.8% of students indicated that it is easy to avoid infection while 21.1% thought it would be difficult. Similarly, 30.2% of staff thought it would be easy to avoid infection while 20.4% of them thought it would be difficult.

**Table 2.** Precautionary measures adopted by the participants to fight COVID-19.

| Precautionary measure | HSC Students |  |  | HSC Staff |  |  |
| --- | --- | --- | --- | --- | --- | --- |
|  | Never | Sometimes | Often/All the time | Never | Sometimes | Often/All the time |
| Wearing a face mask |  |  |  |  |  |  |
| *At home* | 524 (88.5) | 47 (7.9) | 21 (3.6) | 119 (73.5) | 29 (17.9) | 14 (8.6) |
| *Indoor (away from home)* | 16 (2.7) | 29 (4.9) | 547 (92.4) | 15 (9.6) | 24 (15.3) | 123 (75.9) |
| *Outdoor* | 45 (7.6) | 137 (23.1) | 410 (69.3) | 5 (3.2) | 25 (15.9) | 132 (81.5) |
| Washing hands regularly | 7 (1.2) | 110 (18.6) | 475 (80.2) | 1 (0.6) | 11 (6.8) | 150 (92.6) |
| Use disinfectants | 19 (3.2) | 173 (29.2) | 400 (67.6) | 4 (2.5) | 38 (23.5) | 120 (74.1) |
| Avoid contact with elderly | 56 (9.5) | 183 (30.9) | 353 (59.6) | 6 (3.7) | 46 (28.4) | 110 (67.9) |
| Cleaning/disinfecting my phone screen | 61 (10.3) | 249 (42.1) | 282 (47.6) | 13 (8.0) | 53 (32.7) | 96 (59.3) |
| Avoid crowded places | 6 (1.0) | 64 (10.8) | 522 (88.2) | 0 (0) | 10 (6.2) | 152 (93.8) |
| Stay at home as much as possible | 9 (1.5) | 68 (11.5) | 515 (87.0) | 1 (0.6) | 15 (9.3) | 146 (91.1) |
| Avoid eating outside of home | 39 (6.6) | 188 (31.8) | 365 (61.7) | 2 (1.2) | 33 (20.4) | 127 (78.4) |
| Avoid shaking hands when greeting others | 14 (2.4) | 99 (16.7) | 479 (80.9) | 2 (1.2) | 10 (6.2) | 150 (92.6) |
| Avoid kissing others when greeting them | 15 (2.5) | 96 (16.2) | 481 (81.3) | 2 (1.2) | 12 (7.4) | 148 (91.4) |
| Get sufficient sleep | 51 (8.6) | 228 (38.5) | 313 (52.9) | 10 (6.2) | 51 (31.5) | 101 (62.4) |
| Persuade people around you to follow the precautionary guidance | 22 (3.7) | 156 (26.4) | 414 (69.9) | 4 (2.5) | 42 (25.9) | 116 (71.6) |
| Follow social distancing guidance | 6 (1.0) | 93 (15.7) | 493 (83.3) | 0 (0) | 16 (9.9) | 146 (90.1) |

### Sources of information

We assessed the level of trust in COVID-19 information sources among the participants using 5 Likert scale (very little trust, little trust, neutral, moderate, and great deal of trust) against 8 sources of information (Table 3). The greatest deal of trust was in official government press release such as that from the local Ministry of Health (58.8% and 53.7% among student and staff participants, respectively), and consultation with healthcare workers (49.7% and 38.9% among student and staff participants, respectively). Moderate trust was TV stations and online sources (Table 3). Interestingly, there was both very little and little trust (ranging between 60% and 80%) in sources of information from celebrities, social media (such as Twitter, Facebook, and WhatsApp), and conversations with family and friends (Table 3).

**Table 3.** Participants’ responses to questions related to sources of Information

| Source | HSC Students |  |  |  |  | HSC Staff |  |  |  |  |
| --- | --- | --- | --- | --- | --- | --- | --- | --- | --- | --- |
|  | Level of trust |  |  |  |  | Level of trust |  |  |  |  |
|  | Very Little | Little | Neutral | Moderate | Great deal | Very Little | Little | Neutral | Moderate | Great deal |
| TV stations | 55 (9.3) | 68 (11.5) | 171 (28.9) | 184 (31.1) | 114 (19.3) | 23 (14.2) | 20 (12.4) | 38 (23.5) | 49 (30.3) | 32 (19.8) |
| Family & Friends | 195 (32.9) | 190 (32.1) | 140 (33.0) | 58 (9.8) | 9 (1.5) | 40 (24.7) | 39 (24.1) | 46 (28.4) | 26 (16.1) | 11 (6.8) |
| Colleagues | 54 (9.1) | 143 (24.2) | 194 (32.8) | 180 (30.4) | 21 (3.6) | 12 (7.4) | 19 (11.7) | 56 (34.6) | 55 (34.0) | 20 (12.4) |
| Healthcare Workers | 4 (0.7) | 5 (0.8) | 57 (9.6) | 232 (39.2) | 294 (49.7) | 0 (0) | 3 (1.9) | 28 (17.3) | 68 (42.0) | 63 (38.9) |
| Online pages | 71 (12.0) | 116 (19.6) | 181 (30.6) | 174 (29.4) | 50 (8.5) | 17 (10.5) | 22 (13.6) | 56 (34.6) | 44 (27.2) | 23 (14.2) |
| Social Media | 230 (38.9) | 160 (27.0) | 136 (23.0) | 57 (9.6) | 9 (1.5) | 64 (39.5) | 44 (27.2) | 37 (22.8) | 13 (8.0) | 4 (2.5) |
| Official Government press release | 15 (2.5) | 20 (3.4) | 55 (9.3) | 159 (26.9) | 343 (58.0) | 4 (2.5) | 12 (7.4) | 20 (12.4) | 39 (24.1) | 87 (53.7) |
| Celebrities | 482 (81.4) | 65 (11.0) | 33 (5.6) | 7 (1.2) | 5 (0.8) | 103 (63.6) | 25 (15.4) | 25 (15.4) | 7 (4.3) | 2 (1.2) |

Small percent of students indicated that they update themselves about COVID-19 multiple times a day (8.4%); whereas, most of students do that either once a day (34.4%) or once a week (40.3%). In comparison, 25.9% of staff update themselves multiple times a day; whereas 43.8% and 25.3% do that once a day and once a week, respectively.

### Policies related to COVID-19

We listed number of questions related to formulation of policies and regulations about COVID-19. Most of the participants (62.6% and 83.3% of students and staff, respectively) follow ‘very much so’ the recommendations issued by the authorities to prevent the spread of SARS-CoV-2. On questions related to taking COVID-19 vaccination if becomes available, 50% of students strongly agreed and an additional 25.8% agreed to taking it. Interest in vaccine uptake was lower among the staff (28.4% and 34.6% strongly agreed or agreed, respectively).

Participants strongly agreed or agreed (72.5% and 19.6% of students as well as 68.5% and 22.2% of staff) that wearing face mask in public should be obligatory. On the role of government, 67.4% of students strongly agreed and 23.1% agreed on enforcement of quarantine of contacts of COVID-19 cases (compared to 45.7% and 38.2%, respectively among staff). Moreover, 81.4% strongly agreed and 14.4% agreed that there should be a legal punishment for those who know they are infected but continue to socialize with others without wearing protective equipment; whereas, lower percentage of staff strongly agreed or agreed (69.1% and 22.8%, respectively). Furthermore, 55.4% strongly agreed or agreed (i.e., 27.4%) that legal punishment should be taken against those who share/spread falsified information about COVID-19 without verifying the source (compared to 46.3% and 33.3% among staff).

On COVID-19 testing policies, 45.1% and 29.6% strongly agreed or agreed that more COVID-19 testing should be carried out, which is similar to the responses by the staff (40.1% and 30.9%, respectively). Among students, 32.5% and 41.0% strongly agreed or agreed that people should have more information about COVID-19 cases in Kuwait since what is being currently reported has minimal information (i.e., number of new cases, number active and recovered cases, number of PCR tests, number of cases in ICU, and number of deaths). These figures were very similar to the responses by the staff (i.e., 32.8% and 37.7% strongly agreed or agreed). Figure 2 shows the responses about what type of information the participants would like to know about SARS-CoV-2 infectives.

**Figure 2.**
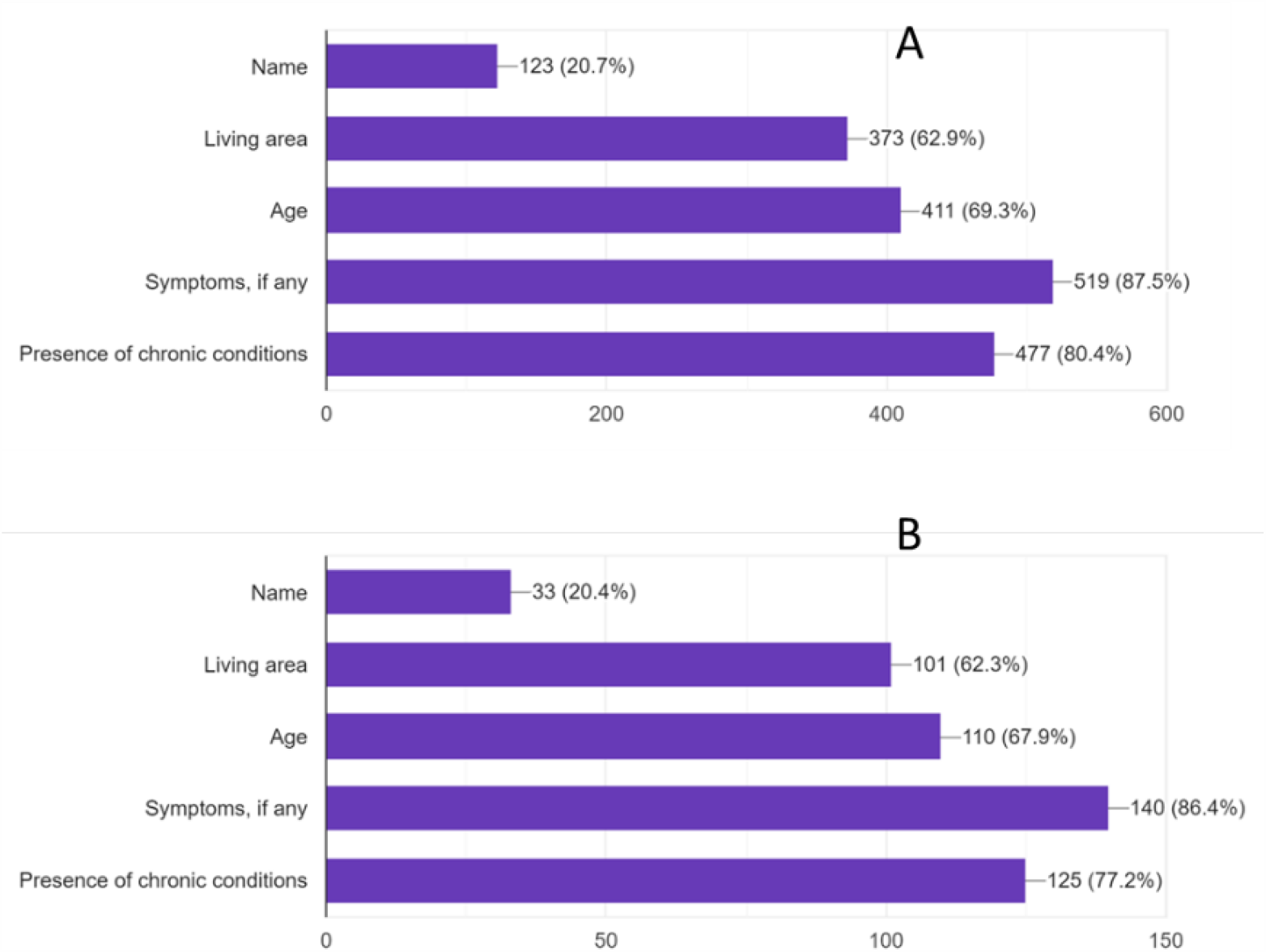
Percentages of participants response to the type of information you would like to know about those infected with SARS-CoV-2. A: students and B: staff.

We asked the participants about their opinion about resuming the university classes on-campus (in person) at the start of the semester in August 2020. Approximately 43% of students and 46% of staff indicated that they are very worried. In addition, 35% of students and 20% of staff indicated that they are slightly worried. Regarding the option of resuming teaching online/distance learning, 75.4% of students and 84% of staff agreed with this option.

### Social Stigma

We asked questions to assess social stigma related to SARS-CoV-2 infection. Interestingly, in response to the question “If anyone you know personally got infected with SARS-Cov-2, what would be your reaction?”, approximately 68% of student and staff participants responded that they would advise the infected to seek medical care immediately; whereas about 29% of both participant groups indicated they would advise to wait for the symptoms to appear before seeking medical care. Interestingly, on the question “Do you think COVID-19 patients did something wrong and consequently got infected”, 12.8% and 18.9% of students strongly disagreed or disagreed; however, 51.8% of them were neutral about it. For the same question, higher proportion of staff 35.2% and 27.8% strongly disagreed or disagreed, compared to 27.2% were neutral about it.

On the question “If you knew a person (e.g., a relative or a friend) has COVID-19, what would you do” with multiple answers permitted, most of the responses by students and staff indicated that they will avoid contact with infected person (83.3% and 80.9%) but understand his/her condition (81.1% and 77.2%). Only 1% or less indicated they would blame the person for getting infected with the virus, but 29% would tell others about the infected person. Moreover, around 20% of students and staff thought that infection with COVID-19 would lead to social rejection; however nearly 75% of students and staff thought COVID-19 would not affect the person’s medical care coverage, job security, finding a job, or getting married. Finally, participants (82.8% of students and 84.6% of staff) indicated that raising awareness about COVID-19 disease will reduce the unjustified stigma toward COVID-19 patients.

## Discussion

Our study focused on COVID-19 awareness and attitude among Kuwait University Health Science students and staff. The university was impacted by the country control measures that included complete closure of its facilities between March – August 2020, followed by partial reopening for staff but continues to offer distance learning for students. Many countries worldwide have gone through school closure and some continue to have it in effect [7]. Our findings indicated that students and staff of HSC in Kuwait University followed precautionary measures to control spread of COVID-19, understood the risk of the virus transmission, sought reliable source of information about COVID-19, and were willing to help those infected with the virus.

COVID-19 epidemic in Kuwait started on February 28, 2020 with imported cases from Iran followed by widespread community transmission that started around March 30, 2020 and still in effect. We revealed that a small percent (1-3%) of the participants reported that have been/currently infected with SARS-CoV-2; however, a larger percent of the participants (>50%) reported knowing someone infected within their immediate social environment indicating a large community transmission in Kuwait. During the study period (May to June 2020), COVID-19 incidence per 100,000 people ranged between 20 – 25 daily new cases with positivity rate between 15 – 20% [6]. Interestingly, self-reported chronic condition among student was high especially asthma (an underlying condition for COVID-19 severe complications). One study reported that the prevalence of asthma among students (age 18 – 24 years) at Kuwait University was 11.9% (n= 1135) [12].

Most of students and staff indicated that they believe that probability of getting infected was low and if they got infected, they likely will not develop severe disease. While this finding was anticipated from young adults (i.e., students), it is less expected from staff especially among those older than 55 years of age [13]. Most of students and staff disagreed that when someone is infected with the virus will likely get admitted to hospital, and if admitted will likely need ICU bed, and then likely to die. This indicates our participants awareness of COVID-19 complication and disease progression among those infected that is similar to what have been reported in the literature [14, 15]. More than 50% of the participants expressed that they will immediately seek medical care if they tested positive for COVID-19 and follow the governmental instructions. This finding is different than what the international health organizations such as CDC recommend [16, 17]. For instance, CDC recommend to stay home even if you test positive, isolate yourself from others, and seek medical care immediately when you have emergency warning signs such as trouble breathing and/or persistent pain or pressure in the chest [16]. Therefore, health education is needed to promote the recommended measures for those who test positive or negative for SARS-CoV-2 or come in contact with a positive case.

Participants followed most of the recommended precautionary guidelines by the WHO [17]. Mask wearing, handwashing, social distancing was common among the participants. Since they represent a particular segment of the community, this population is more likely to adhere to guidelines compare to the general public. Similar findings have been observed among health science and medical students in other countries [11, 18, 19]. Some of the participants indicated their own infection probability is high while others indicated as low. The risk of infection and risk of transmission is multi-factorial depending on host factors, factors related to the virus, and the environment that connects the virus and the host together [20, 21].

Trust in sources of information among participants was mostly in official government sources and information obtained from healthcare workers. Less trust was in information obtained from social media or family and friends. This may reflect the education level of participants and their initiative in seeking information regularly as well as due to the regular dissemination of information by the government communication channel. Similar findings were reported in a study from United States where most of general public participants trusted governmental source (CDC and FDA) compared to private TV networks and social media [22]. Other studies showed similar trend among health science students in Turkey and Iran [23, 24]; whereas, another study from Jordan reported more trust in social media sources among medical students compared to Ministry of Health and international organization such as WHO [25].

The majority of participants indicated that they will take COVID-19 vaccine if becomes available. This is important findings in preparation of vaccination planning and distribution to reach the local herd immunity threshold. To prevent spread of the virus, vast majority of participants indicated that mask wearing should be mandatory with legal punishment for those who do not wear it in public. At the time of this study, there was no state law that fine people for not wearing mask in public. On the other hand, participants indicated that spreading false information or people who know that they are infected and still socialize with others should receive legal punishment. This suggest that participants are aware of the danger of misinformation and irresponsible behavior in spreading the disease.

Social stigma has been reported to have impact on people response to disease control and prevention measures [26-29]. Many participants were not sure if the reason for people getting infected with COVID-19 was due to something they did wrong. Furthermore, participants indicated that they will not blame the person for getting infected. However, participants would avoid contact with infected person, but try to help them which is an approach needed to control disease transmission in the community. Social rejection of patients was a concern shared by the participants. Participants agreed that raising awareness about COVID-19 will lead to reduce unjustified social stigma toward patients as indicated in other studies [27, 28].

This study has several limitations. First, it was based on a convenient sample of students and staff who filled the survey online. The online platform was used due to the pandemic mitigation strategies implemented in Kuwait as well as worldwide. Second, male student participation was low. This limits the generatability of the results to all students. However, the female student population represents about three-fourth of the total student body at HSC. Third, the study assessed the self-reported perception, attitude and awareness. However, this has been the trend among several similar studies in the literature.

## Conclusions

This study presented a baseline estimate of COVID-19 perception and awareness among health science students and staff in Kuwait. It is among the first developed in this special population and provides evidence that more work is needed to raise knowledge and awareness among students in the health science fields. While the span of this epidemic is not clear, health sciences e students and staff may find themselves at the frontlines dealing with patients, especially in the winter season.,..

Universities with similar settings can focus on training their health science students investing in raising knowledge and develop training models for students and staff is a critical and should focus on how to deal with COVID-19 infected people, risk and benefit of treatment and vaccine, and follow precautionary measures recommended by reliable sources of information. Without properly equipped human capital and population commitment, the globe will be at higher risk to consequences of COVID-19 epidemic.

## Supporting information

https://www.dropbox.com/s/m0c845pi5unn0fr/Supplementary%20File%20A.pdf?dl=0

## Data Availability

The datasets used and/or analyzed during the current study are available from the corresponding author on reasonable request

## Ethics approval and consent to participate

All participants were provided with informed consent statement before starting the questionnaire. Agreeing to informed consent prior to starting the questionnaire was mandatory. Moreover, the participants were informed that they have the option to stop answering the questions through the process of completing the questionnaire. The research study was approved by the Ethical Committee of the Health Science Center at Kuwait University (2020/0622). Confidentiality of information was also respected by not asking names and the data was stored with limited access.

## Consent for publication

Not applicable

## Authors’ contributions

Authors (Alali, AlFouzan, and Aljimi) contributed to the study design and development of the questionnaire; Alali, AlFouzan, Aljimi and Al-Tawalah contributed to validation of the questionnaire and its distribution to students and staff; Kheirallah and Yimer contributed to the scientific evaluation of the study design and the questionnaire validity; and Alali, AlFouzan, and Aljimi contributed to the manuscript writeup. All authors read and approved the final manuscript.

## Acknowledgements

The authors would like to thank all of the participants who consented willingly and enrolled in the study voluntarily.

