## Supplementary material for "Perception and awareness of COVID-19 among health science students and staff of Kuwait University: An online cross-sectional study": https://www.dropbox.com/s/m0c845pi5unn0fr/Supplementary%20File%20A.pdf?dl=0

Dear Participants

You are invited to participate in this survey so we can assess the level of knowledge and awareness on topics related to social stigma, source of information, and control and prevention of SARS-CoV-2 (the cause of COVID-19). This survey has been approved by the Ethical Committee of Kuwait University Health Science Center.

Filling the survey is completely voluntary. The survey is designed to identify knowledge gaps in control and prevention of the virus and collect information regarding social stigma and policies related to COVID-19. This survey findings can be communicated back to you.

The survey is composed of 39 questions in six sections. It takes approximately 12 minutes to fill out. We highly appreciate you taking the time to complete this survey.

**Consent of participation:**

** I consent to participate in this survey knowing that my information WILL NOT be used for reasons other than scientific research and it will be confidential.

**Section 1: Socio-demographic Characteristics**

1. What is your gender?
    a. Male
    b. Female

2. Which Faculty are you enrolled in?
    a. Faculty of Medicine
    b. Faculty of Dentistry
    c. Faculty of Pharmacy
    d. Faculty of Public Health
    e. Faculty of Allied Health
3. What is your year of studying?
    a. [drop menu] 1-7 years

4. Do you have any chronic illness?
    a. Yes
        Please specify ______________________
    b. No
    c. Do not know

**Section 2: Risk and knowledge**

1. Are you, or have you been, infected with SARS-CoV-2 (the virus that causes COVID-19)?
    a. Yes, tested and the result was positive
    b. Yes, suspected but not confirmed by a test

   c. No, tested and the result was negative
   d. No
   e. I don't know

2. Do you know people in your immediate social environment (e.g., relatives or friends) who are or have been infected with SARS-CoV-2?
   a. Yes, confirmed
   b. Yes, suspected but not confirmed by a test
   c. No, tested and the result was negative
   d. No
   e. I don't know

3. How would you rate your knowledge level on how to prevent spread of SARS-CoV-2?
   a. Very poor knowledge
   b. Fair knowledge
   c. Moderate knowledge
   d. Good knowledge
   e. Very good knowledge

4. Hot summer weather will slow down/stop the transmission of SARS-CoV-2:
   a. Strongly disagree
   b. Disagree
   c. Neutral
   d. Agree
   e. Strongly agree

5. What do you consider to be your own probability of getting infected with SARS-CoV-2?
   a. Extremely unlikely
   b. Unlikely
   c. Neutral
   d. Likely
   e. Extremely likely

6. How severe would contracting SARS-CoV-2 be for you (how seriously ill do you think you will be)?
   a. Not severe
   b. Slightly severe
   c. Neutral
   d. Moderately Severe
   e. Very severe

7. If you ever suspected that you are infected with SARS-CoV-2, would you:
   a. Go immediately and get tested and follow the government isolation instructions
   b. Wait for few days until symptoms worsen before getting tested

   c. Wait for few days until symptoms worsen before seeking medical care
   d. Stay at home while taking precautionary measures not to spread the virus to others
   e. I don't believe COVID-19 is a serious disease and can be dealt with like seasonal flu

8. Which of the following statements is correct (Select one statement only)?
   a. There is a drug to treat SARS-CoV-2
   b. There is a vaccine for SARS-CoV-2
   c. There is both a drug for the treatment and a vaccine for SARS-CoV-2
   d. There is currently no drug treatment or vaccine for SARS-CoV-2
   e. I don't know

9. Majority of SARS-CoV-2 infected people need admission to a hospital:
   a. Strongly disagree
   b. Disagree
   c. Neutral
   d. Agree
   e. Strongly agree

10. Majority of SARS-CoV-2 patients who are admitted to the hospital require receiving care in the intensive care unit (ICU):
   a. Strongly disagree
   b. Disagree
   c. Neutral
   d. Agree
   e. Strongly agree

11. Majority of SARS-CoV-2 patients who are admitted to the ICU will eventually die
   a. Strongly agree
   b. Agree
   c. Neutral
   d. Disagree
   e. Strongly disagree

**Section 3: Preparedness and prevention:**

1. For me avoiding an infection with the SARS-CoV-2 in the current situation is…
   a. Extremely difficult
   b. Difficult
   c. Neutral
   d. Easy
   e. Extremely easy

2. Which of the following measures have you taken to avoid getting infected with SARS-CoV-2?

| | Never | Sometimes | Often/all the time |
|---|---|---|---|
| Wearing face masks | | | |

| |
|---|
| Washing hands regularly |
| Using disinfectants. |
| Paying more attention to personal hygiene |
| Avoiding contact with elderly |
| Cleaning/disinfecting my phone (screen) |
| Avoiding crowded areas |
| Staying at home as much as possible |
| Avoiding eating outside |
| Avoiding shaking hands when greeting others |
| Avoiding kissing others when greeting them |
| Getting enough sleep |
| Closely monitoring personal physical health. |
| Closely monitoring the physical health of the people around you. |
| Persuading people around you to follow the precautionary guidance. |
| Following social distancing guidance |

3. I follow the recommendations issued by the authorities in Kuwait to prevent spread of SARS-CoV-2.
    a. Not at all
    b. Slightly I do
    c. Neutral
    d. Moderately I do
    e. Very much so

4. What is your opinion regarding the Government's plan of resuming studying in universities in August 2020?
    a. Agree

b. Disagree
c. I don't know

5. What is your opinion in resuming teaching via online /distant learning at Health Science Center faculties?
    a. Agree
    b. disagree
    c. I don't know

6. Please choose the statement that represents your opinion regarding resuming teaching at the university in August 2020:
    a. I am worried about going back to school
    b. I am slightly worried
    c. I am not worried at all about resuming school
    d. I don't know

**Section 4: Information**

1. How much do you trust the following sources of information in their reporting about COVID-19?

| | Very little trust | Little trust | Neutral trust | Moderate trust | A great deal of trust |
|---|---|---|---|---|---|
| Television stations | | | | | |
| Conversations with family and friends | | | | | |
| Conversations with colleagues | | | | | |
| Consultation with health care workers | | | | | |
| Websites or online news pages | | | | | |
| Social media (e.g. Facebook, Twitter, YouTube, WhatsApp) | | | | | |
| Official government press releases | | | | | |
| Celebrities and social media influencers | | | | | |

2. How often do you update yourself about COVID-19?
    a. Never
    b. Once a month
    c. Once a week
    d. Once a day
    e. Multiple time a day

3. SARS-CoV-2 virus was manufactured in a laboratory:
    a. Strongly agree
    b. Agree
    c. Neutral
    d. Disagree
    e. Strongly disagree

**Section 5. Policies**

What is your opinion regarding the following statements?

1. SARS-CoV-2 vaccine will be available for use within:
    a. Six months
    b. One year
    c. Two years
    d. Never
    e. Do not know

2. If a vaccine becomes available and is recommended for me, I will get it.
    a. Strongly disagree
    b. Disagree
    c. Neutral
    d. Agree
    e. Strongly agree

3. If a treatment becomes available and is recommended for me, I will get it.
    a. Strongly disagree
    b. Disagree
    c. Neutral
    d. Agree
    e. Strongly agree

4. The government should be allowed to force people into self-isolation if they have been in contact with a person who was infected
    a. Strongly disagree
    b. Disagree

c. Neutral
d. Agree
e. Strongly agree

5. Wearing a face mask in public areas should be obligatory
    a. Strongly disagree
    b. Disagree
    c. Neutral
    d. Agree
    e. Strongly agree

6. There should be a legal punishment for those who know they are SARS-CoV-2 infected and continue socializing with others without personal protective equipment (PPE).
    a. Strongly disagree
    b. Disagree
    c. Neutral
    d. Agree
    e. Strongly agree

7. There should be a legal punishment for those who share/spread falsified information about COVID-19 without verifying the source.
    a. Strongly disagree
    b. Disagree
    c. Neutral
    d. Agree
    e. Strongly agree

8. More tests for SARS-CoV-2 infection should be carried out in the population
    a. Strongly disagree
    b. Disagree
    c. Neutral
    d. Agree
    e. Strongly agree

**Section 6. Social Stigma**

1. If anyone you know personally got infected with SARS-Cov-2, what would be your reaction?
    a. Advise them to seek medical care immediately
    b. Advise them to wait for symptoms to appear before seeking medical care
    c. Advise them to keep it secret because of social stigma
    d. Advise them NOT to seek medical care because of crowded hospitals or other reasons
    e. I don't believe COVID-19 is a serious disease and can be dealt with like seasonal flu

2. If you knew a person (e.g., a relative or a friend) has COVID-19, what would you do? (Choose all that applies):

a. Avoid contact with him/her
b. Tell others about him/her
c. Fully blame him/her for getting infected
d. Partially blame him/her for getting infected
e. Understand his/her condition and try to help

3. Do you think COVID-19 patients did something wrong and consequently got infected?
    a. Strongly disagree
    b. Disagree
    c. Neutral
    d. Agree
    e. Strongly agree

4. What type of information you would like to know about those infected with SARS-CoV-2? (Choose all that applies)
    a. Name
    b. Living area
    c. Age
    d. Symptoms, if any
    e. Presence of chronic conditions

5. Do you think people should have more information about COVID-19 cases in Kuwait?
    a. Strongly disagree
    b. Disagree
    c. Neutral
    d. Agree
    e. Strongly agree

6. Do you think if a person has COVID-19, this will lead to: (Choose all that applies)
    a. Being socially rejected
    b. Being denied medical care
    c. Lose his/her job
    d. Harder to find a job
    e. Harder to get married

7. How can one reduce the unjustified stigma toward COVID-19 patients? (Choose all that apply)
    a. Increase awareness about COVID-19
    b. Increase awareness about the virus mode of transmission
    c. Clarify that a person can get infected despite taking all prevention measures
    d. Clarify that a person can be infectious to others but shows no symptoms (asymptomatic transmission)
    e. Increase awareness about not spreading false information

Thank you very much! Your participation provides valuable insights for all of us to react appropriately in the current COVID-19 situation and to reach all citizens with useful information in a timely manner.
